# Simplified model of the number of Covid-19 patients in the ICU: update April 6, 2020

**DOI:** 10.1101/2020.04.07.20056226

**Authors:** A. Cividjian, F Wallet, C. Guichon, O. Martin, S. Couray-Targe, T Rimmelé, PF Wey

## Abstract

**INTRODUCTION:** Predicting the number of Covid-19 patients in the Intensive Care Units (ICU) could be useful to avoid the breaking point. We attempted to deduce a formula in order to model the number of the ICU patients in France from the official data and patient turnover in the ICU.

**METHODS:** The Covid-19 ICU patient turnover was calculated using a recurrence relation from the internal data provided by Hospices Civils de Lyon. The number of new Covid-19 cases detected daily was modelized to fit with the last known data in France and extrapolated for the coming days using two scenarios following the existing data in China (best scenario) and Italy (worst scenario). The number of daily admissions in ICU was calculated as the sum of 13.7% of the new Covid-19 cases detected on a given day and 7.8% of the average of the total new Covid-19 cases recorded in the last week. Approximately 39.7% of patients admitted to the ICU were non-intubated with an average ICU length of stay of 4 days. Conversely, 60.3% of patients were intubated and for those who died among them (14.44%) the ICU length of stay was of 4 days for 78.3% of them and of 15 days for 21.7% of them. For the intubated patients that were discharged alive, the ICU length of stay was of 6 days for 44.4% of them and of 20 days for 55.6% of them.

**RESULTS:** We predict a peak of 7072 – 8043 patients for the overall French territory.

**CONCUSION:** Despite a simplified mathematical model, the strength of our study is a narrow possible range of predicted total number of ICU patients.

## INTRODUCTION

Most of European national health care services faced an historic and never observed acute issue due to SARS-Cov2 (Covid19) pandemic situation. Several Intensive Care Units (ICU) in Italy, Spain, France reached the breaking point, the strain on ICU free beds leading to patient evacuation and ethical considerations.

A previous study (1) predicted during the early stages of the pandemic that between 2,500 and 25,000 patients could require an ICU stay in France. However, this predicted range depending on an unknown value of R0 is very large, and a narrower prediction range could be useful. In the present stage of the pandemic, more information is available about the pathology. The aim of this study is to perform a simplified mathematical model predicting the number of ICU beds available for Covid-19 patients from the observed ICU patient turnover for this precise pathology rather from the R0 value. As these patients’ length of stay in the ICU may exceed 21 days, there is a cumulative effect that may delay by several days the peak of the number of patients in the ICU as compared to the epidemic peak. We attempted to deduce a formula in order to model the number of the ICU patients in France from the official number of confirmed cases of Covid-19 and from the Covid-19 patient turnover in the ICU.

## METHODS

The Covid-19 ICU patient turnover was calculated using a recurrence relation that amounts to the sum between the number of existing patients and the number of daily admissions from which was subtracted the number of patients discharged from the ICU, dead or alive.

### Number of daily admissions in the ICU

the number of daily admissions was calculated as the sum of a ratio k_d_ of the new Covid-19 cases detected on a given day D0 and a ratio k_w_ of the average of the total new Covid-19 cases recorded in the last week (W0), <u>due to complications of an initially stable</u> patient state. The ratios k_d_ and k_w_ after the start of containment were estimated from the internal data provided by Hospices Civils de Lyon (3): k_d_ = 0.137 and k_w_ = 0.078.

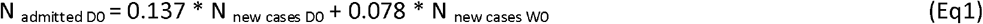

Before the containment, the ratios k_d_ and k_w_ were lower (k_d_ = 0.03 and k_w_ = 0.015), because all the suspicious Covid-19 cases were tested.

The number of new Covid-19 cases detected daily was modelized to fit with the last known data in France and extrapolated for the coming days using two scenarios following the existing data in China and Italy (2). This extrapolation was performed by a linear transformation of the existing curve of the total number of detected Covid-19 cases in China and Italy as follows: the same value at the inflection point (3-point average centered on the inflection point) and the same derivative after the inflection point (average of 3 consecutive derivatives after the inflection point) as the curve of the total number of detected Covid-19 cases in France (3) was required.

### ICU length of stay

The length of stay of the Covid-19 patients in the ICU depends on the severity of those patients’ condition, which could be stratified as a function of the ventilation mode. Indeed, following internal data from Hospices Civils de Lyon (4), approximately 39.7% of patients admitted to the ICU were non-intubated (nasal cannula O2, non-invasive ventilation e.g. Optiflow) with an average ICU length of stay of mean ± standard deviation (SD) 3.74 ± 1.76 days for those who were discharged alive and 3.92 ± 3.29 days for those who died. The ICU length of stay was rounded to 4 days for all the non-intubated patients. Conversely, 60.3% of patients were intubated and for those who died among them (14.44%) the ICU length of stay was of 4.33 ± 2.3 days (rounded to 4 days) for 78.3% of them and of 14.6 ± 3.9 days (rounded to 15 days) for 21.7% of them. For the intubated patients that were discharged alive, the ICU length of stay was of 5.5 ± 3.32 days (rounded to 6 days) for 44.4% of them with less severe respiratory failure, and of 20.4 ± 3.29 days (rounded to 20 days) for 55.6% of them with more severe respiratory failure and/or associated complications. Thus, the number of patients discharged the day D0 could be approximated as follows:

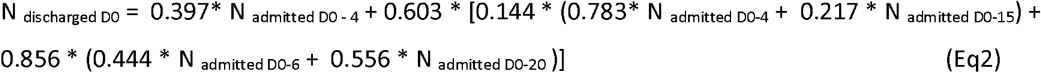

Thus, the cumulated number of patients in the ICU the day D0 is:

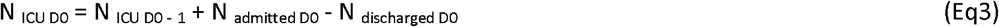

The number of patients admitted and discharged on day D0 was calculated using respectively (Eq1) and (Eq2) with an Excel (Microsoft) spreadsheet.

## RESULTS

The resulting extrapolations of confirmed Covid-19 cases following the Chinese and Italian scenarios are reported in the Figure 1.

**Figure 1.**
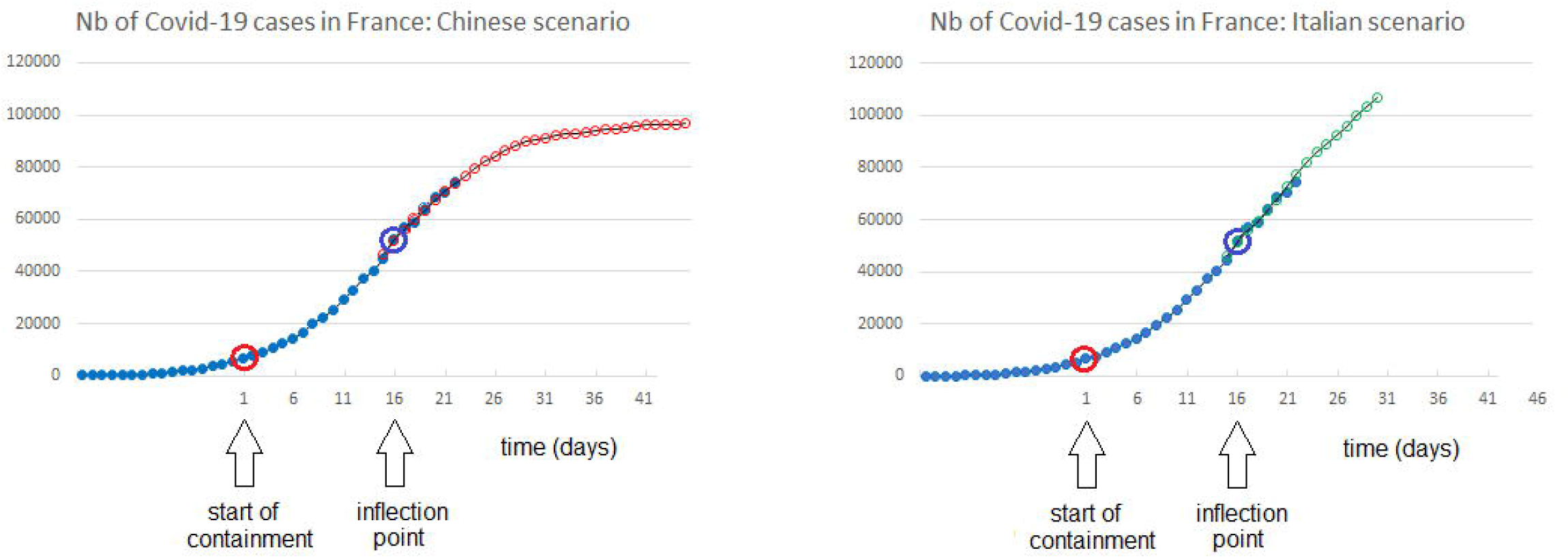
Extrapolation of the total number of confirmed Covid-19 cases in France (blue solid points) following Chinese (left, red open points) and Italian (right, green open points) scenarios. Red circle: the start of effective containment in France (March 17). Blue circle: supposed inflexion point (March 31).

The resulting extrapolations of the number of patients present in the ICU following the Chinese and Italian scenarios are reported in Figure 2.

**Figure 2.**
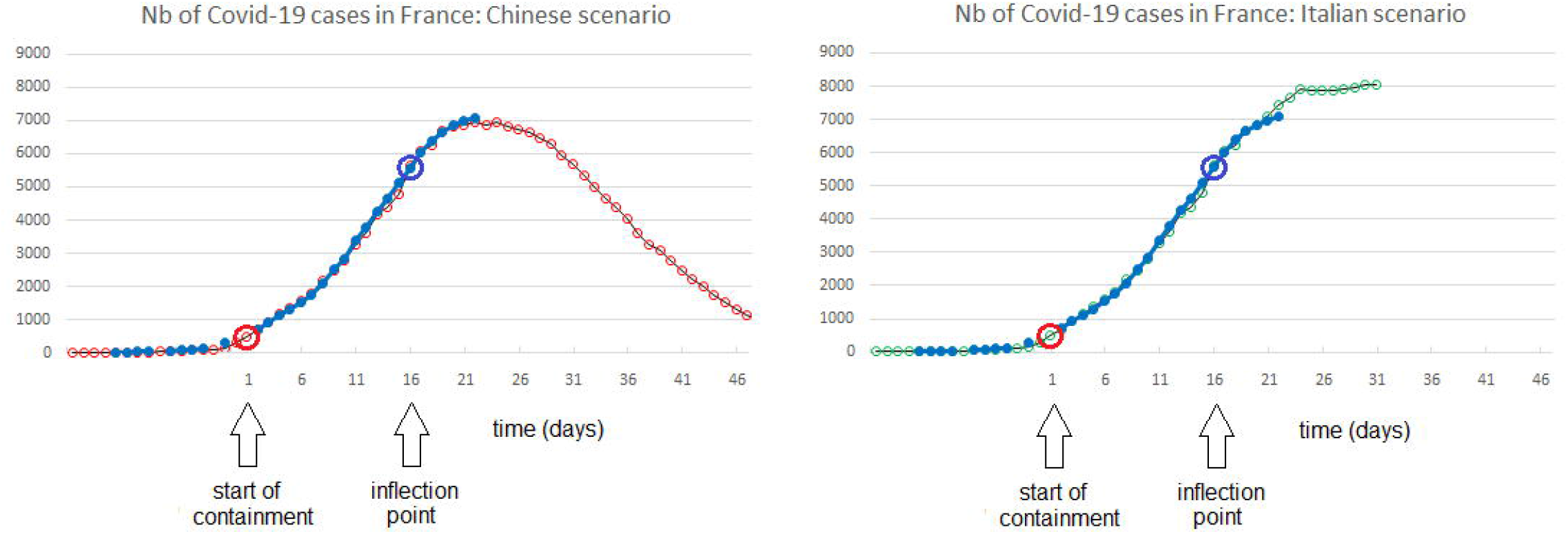
Extrapolation of the number of Covid-19 patients present in the ICU in France (blue solid points) following the Chinese (left, red open points) and Italian (right, green open points) scenarios. Red circle: the start of effective containment in France (March 17). Blue circle: supposed inflexion point (March 31).

## DISCUSSION

This mathematical model presents several limitations. First, the spread of Covid-19 is not homogeneous in France, with a higher density in the Eastern part of France and around Paris. The presented model uses all the pooled data irrespective of the widespread. Second, all the proportions and coefficients used in the model can vary in time and between hospitals. Third, age and patient comorbidities were not considered. The variability of all these factors should be considered for a more accurate model.

The formula used in the model is the same for both scenarios, the only difference being the curve of the new Covid-19 cases. The Chinese and the Italian scenarios were taken as references because the former corresponds to an ideal case with quasi-absolute containment, and the latter corresponds to one of the highest known cases of prevalence with containment conditions similar to the French ones. Thus, the strength of our study is a narrow possible range of predicted total number of ICU patients, namely a peak of 7072 – 8043 patients for the overall French territory. Such a prediction could help to avoid the ICU bed capacity reaching breaking point. Mixing our method with the already published methods such as (1) could yield more precise results.

A last point, is that after the start of containment, only the severe cases of Covid-19 have been tested in France and consequently the ratios k_d_ = 13.7% and k_w_ = 7.8% estimated from the Hospices Civils de Lyon (4) were higher than the true prevalence of severe Covid-19 cases requiring admission in ICU.

## CONCLUSION

Following the presented mathematical model, two scenarios are possible in the following days (April 5-8): in the case of a Chinese scenario the number of patients in the ICU will remain on a plateau, whereas in the case of an Italian scenario the number of patients in the ICU could still increase by 13.7%.

## Data Availability

Excel formulas are available on request

## ACKNOWLEDGMENTS

Dominique Delmaire from University Lyon II reviewed the manuscript.

